# Are aortic biomechanical properties early markers of dilatation in patients with Marfan Syndrome? – A Systematic Review and Meta-Analysis

**DOI:** 10.1101/2024.01.05.24300824

**Authors:** Claire Rosnel, Raphael Sivera, Elena Cervi, Mark Danton, Silvia Schievano, Claudio Capelli, Ankush Aggarwal

## Abstract

**Background:** Although the stiffness of tissue is known to play an important role in aortic dilatation, the current guidelines for offering a preventative aortic surgery in patients with Marfan syndrome rely solely on the aortic diameter. In this systematic review and meta-analysis, we analyze and compare literature on in-vivo aortic stiffness measurements in Marfan patients. Our aim is to assess the potential of these measurements as early indicators of aortic dilatation.

**Methods:** Following the PRISMA guidelines, we collected literature on diameter and three in-vivo stiffness measures: Pulse Wave Velocity (PWV), *β*-stiffness index and Distensibility, at five different aortic locations in patients with Marfan syndrome. Reported results were reviewed and compared against each other. For meta-analysis, an augmented dataset was created by combining extracted data from the reviewed literature. Regression with respect to age and statistical comparison were performed on the augmented dataset for all three measures at five different locations.

**Results:** 30 articles reporting data from 1925 patients with Marfan and 836 patients without Marfan were reviewed. PWV was found to be statistically higher in Marfan at most aortic locations, but only when the aorta is already dilated. Distensibility was found to be lower at all aortic locations even in non-dilated aortas, and its decrease has been associated with higher chances of developing aortic dilatation. *β*-stiffness index was higher in Marfan patients and was positively correlated with the rate of aortic dilatation, emphasizing its role as a valuable indicator. In our meta-analysis based on a total 1197 datapoints, diameter was found to be higher only at the root (*p <* 0.001). All stiffness measures showed a significant variation with age. PWV at the root and carotid-femoral region was not statistically different (*p* = 0.62 and *p* = 0.14 respectively), but was positively correlated with age at all locations. Distensibility and *β*-stiffness index were different in Marfan patients at all locations, and the difference was more pronounced after accounting for age-related variation.

**Conclusion:** Based on the results in the literature, *β*-stiffness index and distensibility emerge as the best predictors of future aortic dilatation. Our meta-analysis quantifies age-related changes in aortic stiffness and highlights the importance of accounting for age in comparing these measurements. Missing diameter values in the literature limited our analysis. Further analysis based on combined aortic stiffness and diameter criteria is recommended to evaluate aortic disease in a comprehensive way and assist clinical decisions for prophylactic surgery.

## 1 Introduction

Marfan Syndrome (MFS) is a heritable connective tissue disorder caused by a mutation of the fibrillin-1 gene (FBN1). The FBN1 mutation increases the fragmentation of elastic fibers in the aortic media, leading to compromised strength and structure of the tissue [1]. As a consequence, MFS exposes patients to higher risks of aortic diseases such as dilatation, dissection and rupture due to stiffening of the vessel’s walls. 50% of undiagnosed and untreated MFS patients die by the age of 40 due to cardiovascular complications [2]. Early diagnosis via cardiovascular imaging is thus crucial for suspected Marfan patients, and close monitoring is essential for confirmed patients. Echocardiograms and Magnetic resonance imaging (MRI) are examples of imaging procedures conducted to evaluate changes in aortic size and expansion rate. The current 2022 ACC/AHA clinical guidelines for preventive surgical intervention is largely based on the aortic root diameter with a threshold at 50mm, or 45mm in patients with increased risks of aortic dissection [3]. Surgery is also recommended when the cross-sectional aortic root area to patient height ratio is greater than 10 cm^2^/m. However, dissection and rupture are known to occur below these thresholds [4, 5, 6], and diameter alone may not fully account for the biomechanical properties of aortic tissue, which are expected to play an important role in aneurysm progression and adverse events. While only diameter is currently used to predict risks of dissection and rupture, aortic stiffness emerges as a predictor of aortic dilatation, offering insights into the probability of adverse events.

Considerable research has focused on investigating how biomechanical properties of the aorta can be early predictors of dilatation using in-vivo measures of aortic stiffness, namely 1) pulse wave velocity (PWV), 2) distensibility, and 3) *β*-stiffness index. Although some results have shown that aortic stiffness measured in-vivo may perform better than diameter assessment to predict aneurysmal growth, no consensus has been established on what stiffness indicator to use, which aortic segment to consider, how it differs from healthy patients, and how the differences evolve with age.

This systematic review of the literature therefore aims at gathering published research studies that report aortic stiffness using PWV, distensibility and *β*-stiffness index in patients with Marfan syndrome and healthy patients. A comprehensive analysis of their correlation with age, and their potential as early indicators of aortic dilatation is conducted. In addition, to overcome variations in individual studies, we aim to create a larger, consolidated dataset from the selected studies and perform a meta-analysis to determine age– and disease-related variations and differences.

## 2 Methods

The following systematic review adheres to the Preferred Reporting Items for Systematic Reviews and Meta-analyses (PRISMA) recommendations and guidance [7]. In this section, the eligibility and search criteria necessary for the identification and selection of relevant published studies are defined. The data extraction and augmentation process, as well as the statistical techniques used to conduct the meta-analysis are also described.

### 2.1 Inclusion criteria, information sources and search strategy

Studies focusing on one or more of the three clinically established aortic stiffness measures – PWV, distensibility and *β*-stiffness index – were selected for patients diagnosed with MFS. Regarding the diagnosis of MFS, the revised Ghent criteria is the most widely accepted since its proposition in 1996 [8]. Therefore, only the studies published between 1996 and September 2022 were included. Cohort studies, cross-sectional studies, case-control studies, and case series were considered, whereas conference abstracts, book chapters, case reports, reviews, editorials, expert opinions and letters were excluded. The review focused on early signs of dilatation, and thus excluded papers investigating severe complications such as dissection and rupture. Articles focusing on the following aspects were also excluded: effect of medication on aortic stiffness, ex-vivo mechanical characterization, cellular scale investigations, and effect of aortic curvature on its mechanical behavior. Additionally, the review was restricted to publications in the English language.

Two electronic databases, PubMed and ScienceDirect, were screened to find publications based on the inclusion criteria. A time filter was applied to encompass research published between 1996 and September 2022. Between May 2022 and September 2022, the databases were searched using the following MeSH terms: ‘Aortic’ AND ‘Stiffness’ AND ‘Marfan’,

### 2.2 Clinical metrics of aortic stiffness

Stiffness refers to the ability of a material to withstand deformation under an applied force. The in-vivo stiffness of arteries is influenced by their geometry and the biomechanical properties of the tissue. In this section, we define the three in-vivo parameters that are used in this review to quantify aortic stiffness.

∎ PWV is defined as the speed of the pressure waveform over a designated portion of a vessel. Higher PWV values indicate a wave that travels faster along the arterial segment, generally a consequence of stiffer tissue. The wave is detected using pressure transducers or Doppler echocardiography, and its travel time, called transit time, is measured by estimating the time of travel of the foot of the wave over a known arterial distance. The pulse wave velocity is therefore calculated as the distance between two chosen points divided by the transit time, 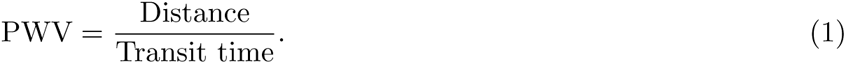
∎ Distensibility (*Dist*) is directly calculated on in-vivo images. It is defined as the relative change in a vessel’s luminal area for a unit pressure increment. Thus, a lower distensibility value indicates stiffer tissue. From in-vivo images, the arterial luminal area at systole and diastole, denoted as *A_s_* and *A_d_* respectively, are measured at a chosen location. Pressure measurements are commonly taken at the brachial artery using a sphygmomanometer cuff, where the systolic pressure (denoted as *P_s_*) and diastolic pressure (denoted as *P_d_*) are measured. Distensibility is thus calculated as 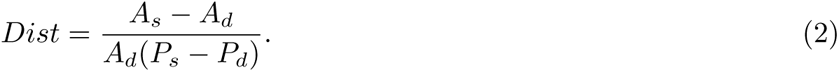
∎ The *β*-stiffness index (*β*-SI) is also derived from in-vivo image-based measurements. Higher values indicate a stiffer tissue, i.e., *β*-SI is inversely correlated to the distensibility. *β*-SI is defined as the logarithm of the pressure ratio to relative change in diameter, and is dimensionless. The diameter in systole and diastole is measured on cross-sectional views of the aorta, and pressure using sphygmomanometer cuff. *β*-SI is calculated 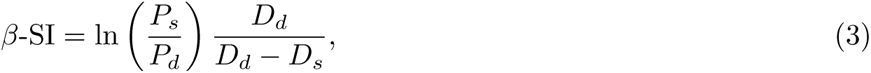 where *P_s_*, *P_d_*, *D_s_* and *D_d_* are the systolic pressure, diastolic pressure, systolic diameter and diastolic diameter, respectively.

### 2.3 Data collection process

The details of the selected papers and their full-text manuscripts were stored in a reference management software (Zotero). Information from these manuscripts was extracted, capturing the following data: publication details, overall aim of the study, study design (prospective, retrospective, multicenter, or longitudinal), cohorts’ size, cohorts’ mean age, imaging modality (Echocardiography (Echo) or Magnetic Resonance Imaging (MRI)), presence of diagnosed aneurysms in the cohort, and the aortic stiffness measure reported at five aortic locations: aortic root, ascending aorta (Aao), aortic arch (Arch), descending aorta (Dao), and carotid-femoral (only for PWV).

Discrepancies in results from the literature may be attributed to the specificity of the cohorts included in each individual paper, particularly factors like age and population size. Additionally, statistical tests conducted on their sampled cohorts might not be fully representative of the broader population. A meta-analysis was therefore sought on the following quantities of interest: PWV, distensibility, *β*-stiffness index, diameter, and age. The aim was to perform a comprehensive statistical analysis by treating the collective results from the literature as one unified dataset, and offering deeper insights than what can be derived from individual papers. The unified dataset creation required extraction of data points from selected manuscripts, but with the following exclusion criteria. For studies investigating several connective tissue disorders, measurements were excluded from the meta-analysis if Marfan data points could not be separated from others. Since the focus of this analysis is on the native biomechanical properties without any effects of surgical intervention, measurements were also rejected if results from patients who underwent an aortic surgical procedure (e.g., PEARS, Bentall procedure) were not separable from the rest of the Marfan cohort. However, data points with unknown surgical status were included under the assumption that if surgery was performed, it would be explicitly mentioned in the respective articles. Patients under medication were included since that they represent a large portion of the diagnosed Marfan population. When studies presented data separately for aneurysmal and non-aneurysmal Marfan aortas, only the non-aneurysmal measurement was selected. For longitudinal studies with several time points reported, only the baseline measurement was extracted. Finally, in papers where results were reported per age range, the mean was calculated and collected. For articles where individual participant data points were available, they were digitized directly from plots in the manuscript using WebPlotDigitizer [9]. In cases where such individual data points were not provided, mean and standard deviation (*SD*) or median and interquartile ranges (IQR) were manually extracted for each quantity.

### 2.4 Data augmentation process

In the literature, various image-based aortic stiffness measures are employed to characterize the mechanics of vascular walls. However, the measures often use different units, leading to inconsistency and lack of standardization, as previously pointed out by Alhalimi et al. [10]. To address this issue, our data augmentation process involved employing standardized formulae and units, as well as conversion equations to transform one aortic stiffness index into another. Specifically, distensibility can be converted into *β*-SI using the relative change in area (*A_s_* − *A_d_*)*/A_d_* = 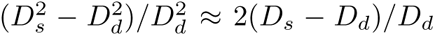 where *A_s_*, *A_d_*, *D_s_*, *D_d_* are systolic area, diastolic area, diameter in systole and diameter in diastole respectively. The conversion of PWV to image-based aortic stiffness measures can be achieved using the Bramwell-Hill equation [11]. The conversion formulae are summarized in Table 1, where *ρ* is the blood density approximated to be 1059 *kg/m*^3^. Mean values of systolic (*P_s_*) and diastolic pressure (*P_d_*) reported in the articles were used for the calculations. It is worth noting that in cases where only the pulse pressure (*P_s_* − *P_d_*) was reported instead of systolic and diastolic pressures individually, the *β*-stiffness index could not be calculated.

**Table 1:**
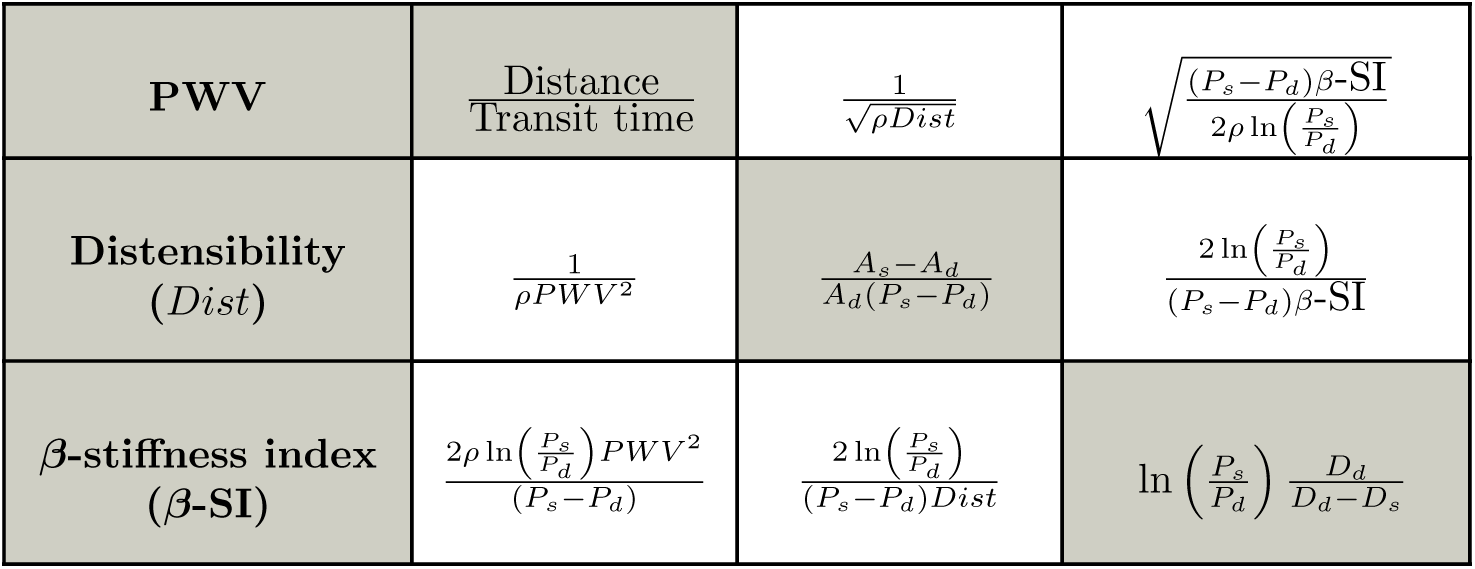
Calculation equations (in gray cells) and conversion equations (in white cells) for the three aortic stiffness measures.

In instances where individual data points were not reported, mean and standard deviation of the quantity were used. However, as noted by Weir et al. [12], when results exhibit a skewed distribution, researchers often report the median and interquartile ranges instead of the mean and variance information. To ensure that such cases were not excluded from the analysis, missing mean and standard deviation values were calculated from the provided median and interquartiles using Wan et al.’s method [13], described as follows. The mean *x*~ can be estimated from median and interquartiles as

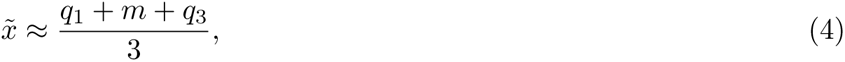

where *q*_1_ and *q*_3_ represent the first and third interquartiles, and *m* denotes the median. The standard deviation *SD* is estimated as

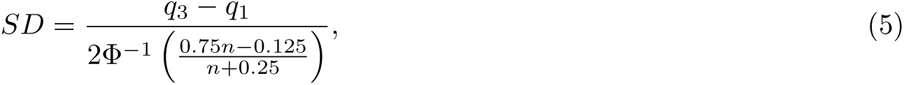

where Φ is the cumulative distribution function of a standard normal distribution and *n* is the size of the cohort.

To summarise, the augmented dataset on which the meta-analysis is conducted consisted of three types of data:

(i) Individual patients’ data points that were directly digitized and collected from plots in the manuscripts.
(ii) Mean and standard deviation values collected from the manuscripts.
(iii) Mean and standard deviation values calculated from the median and interquartiles reported in manuscripts using Wan et al.’s method [13].
(iv) Calculated values, which were generated using the conversion equations from Table 1 to transform one reported aortic stiffness measure into another.

### 2.5 Statistical analysis

In the meta-analysis, comparison tests between Marfan and control and linear regressions with age were performed on the augmented dataset. As stated previously, the dataset is composed of individual datapoints as well as mean and standard deviation values. In order to run statistical tests, all values were expressed as mean and SD, such that the overall means for the Marfan (*i* = *M*) and control (*i* = *C*) cohorts are calculated as

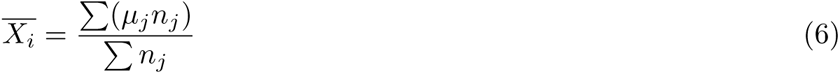

where *µ_j_* and *n_j_* are the mean and size of the cohort in paper *j*, and the corrected sample standard deviations are calculated as

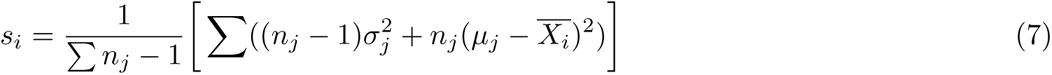

where *σ_j_* is the standard deviation in paper *j*.

Marfan and control were compared using the Welch test, which examines the null hypothesis that two populations have equivalent means. This test is favored over the Student’s *t*-test when the two samples have unequal variances. Bonferroni correction was applied to account for multiple comparisons type two error.

To better understand the independent effect of Marfan syndrome on distensibility, PWV or *β*-SI and to account for age-related effects, linear regressions and projections at age-zero were conducted. To perform the regressions, the normality of the dataset was tested using Shapiro–Wilk test. Due to the lack of consistent normality in the initial data distribution, a logarithmic transformation was applied to the dataset. The association between variables and age were evaluated using the coefficient of determination (*R*^2^). The slope of the linear regression enables us to determine if age-related changes occur at a faster rate in Marfan patients. To investigate whether Marfan patients are born with altered stiffness or if it changes over time, log-values of the three aortic stiffness measures were projected at age-zero using the linear regression. Welch comparison tests were then run on the projected values. The projection allowed us to discern whether statistical differences between the two groups can be found once the age-related variations had been factored out. In all statistical tests, significance was considered at a *p*-value less than 0.05. All analyses were performed using Python and the Scipy library.

## 3 Results

### 3.1 Search results

The flowchart in Figure 1 illustrates the paper selection process following PRISMA guidelines. Initially, 639 published articles were identified, comprising 76 papers from PubMed, 553 from Science Direct, and 10 from reference list hand-searching. After removing duplicates, 616 papers remained, which were then assessed against exclusion/inclusion criteria by examining the title and abstract only. Among them, 572 articles were excluded, primarily for being unrelated to in-vivo measures of aortic stiffness in Marfan diagnosed patients, or for focusing on blood flow patterns. Subsequently, 44 texts were read fully, and 14 were rejected for either not using the revised Ghent criteria for MFS diagnosis [8] or for merging Marfan patients with other tissue disorders. Finally, a total of 30 papers were selected based on the eligibility criteria and search strategy mentioned in Methods section. For the meta analysis, 6 of the 30 articles were excluded for not reporting Marfan data separately from others.

**Figure 1:**
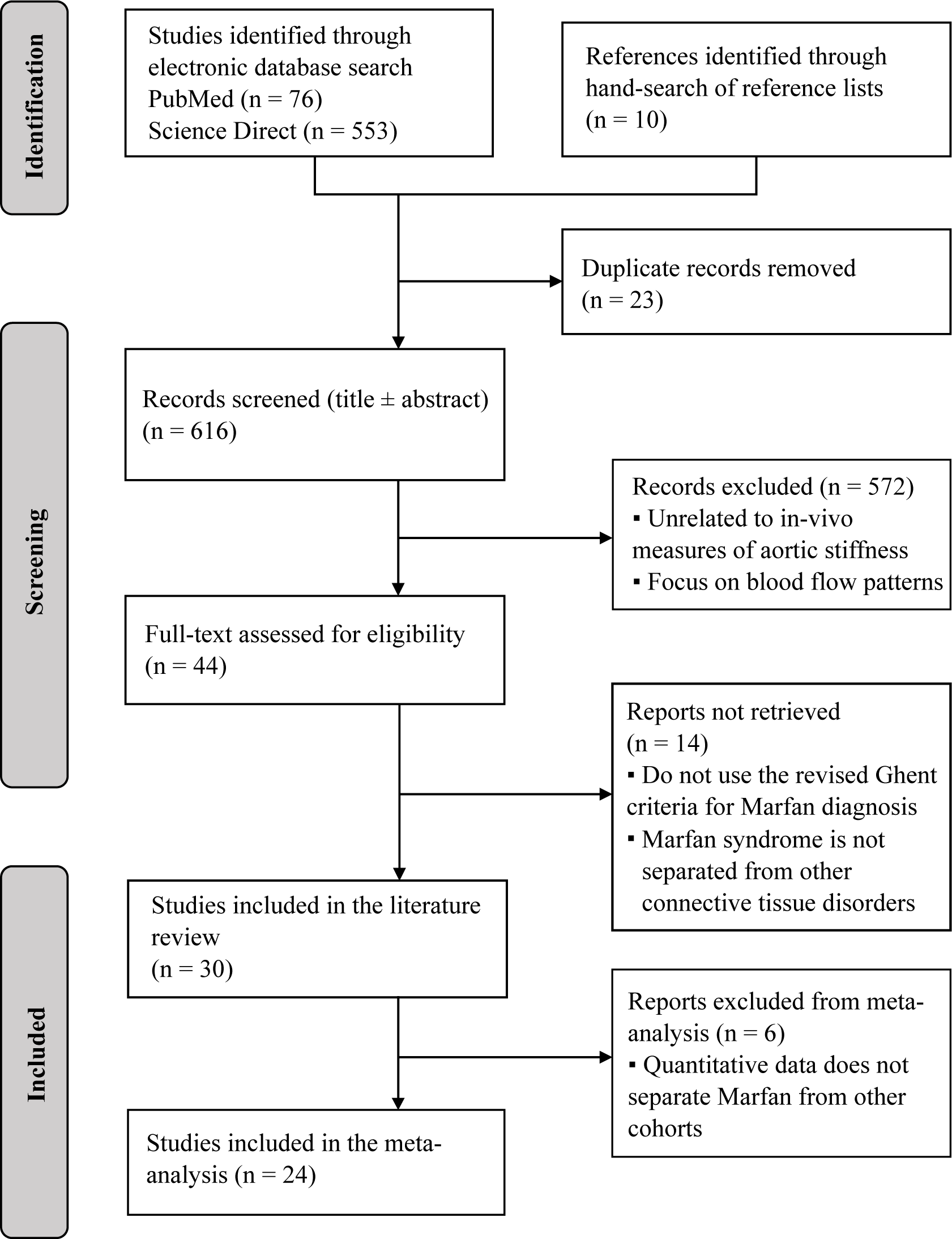
The flow diagram presents the process of inclusion given the eligibility criteria using the PRISMA 2020 guidelines

### 3.2 Characteristics of included studies

The review of the literature offered valuable insights into the assessment of stiffness in patients with Marfan Syndrome, as it encompassed studies from diverse sources published over a span of 20 years in ten different countries. Out of the 30 selected studies, 16 were longitudinal, providing crucial information on how stiffness evolves with age. The remaining 14 were case-control studies, allowing for a quantification of the differences in stiffness properties between individuals with and without Marfan Syndrome.

Of the selected papers, 17 used echocardiography to monitor the size of the aorta and to calculate stiffness measures at various aortic locations, while 13 employed MRI. Only one study by Prakash et al. [14] utilized both imaging modalities: MRI for distensibility and *β*-SI calculation and echocardiography for monitoring the aortic size. Table 2 provides a comprehensive overview of the included papers in the qualitative synthesis, along with their respective main characteristics.

**Table 2:** Main characteristics of the 30 studies included in the systematic review and meta-analysis.

| Author | Country | Marfan cohort: size (mean age) | Control cohort: size (mean age) | Imaging modality | Included in the meta-analysis? | Quantity reported |
| --- | --- | --- | --- | --- | --- | --- |
| Groenink et al. [15] | Netherlands | 78 (31) | 23 (28) | MRI | Yes | PWV, Distensibility |
| Sandor et al. [16] | Canada | 14 (15.7) | 6 (12.3) | Echo | Yes | PWV, β-SI |
| Nollen et al. [17] | Netherlands | 78 (31) | - | MRI | Yes | PWV, Distensibility |
| Baumgartner et al. [18] | Austria | 19 (17.7) | 19 (17.7) | Echo | Yes | Distensibility, β-SI |
| Oosterhof et al. [19] | Netherlands | 78 (31) | 17 (44) | MRI | Yes | PWV |
| Bradley et al. [20] | Canada | 26 (13.14) | 69 (13.14) | Echo | Yes | PWV, β-SI |
| Vitarelli et al. [21] | Italy | 31 (26) | 21 (26) | Echo | Yes | PWV, Distensibility, β-SI |
| Baumgartner et al. [22] | Austria | 46 (17.4) | 46 (17.6) | Echo | Yes | Distensibility, $\beta$ |
| Fattori et al. [23] | Italy | 20 (27.8) | 14 (29) | MRI | Yes | Distensibility |
| Mortensen et al. [24] | Germany | 50 (32) | - | Echo | Yes | PWV |
| Kiotsekoglou et al. [25] | U.K | 31 (31) | 31 (33) | Echo | Yes | PWV, β-SI |
| Westenberg et al. [26] | Netherlands | 25 (36) | 25 (36) | MRI | Yes | PWV |
| Wit et al. [27] | Australia | 55 (40.5) | 69 (41.35) | Echo | Yes | PWV, distensibility, β-SI |
| Kröner et al. [28] | Netherlands | 21 (36) | 26 (30) | MRI | Yes | PWV |
| Teixido-Tura et al. [29] | Spain | 80 (32) | 36 (35.2) | MRI | Yes | PWV, Distensibility |
| Prakash et al. [14] | U.S | 45 (27) | - | Echo, MRI | No | Distensibility, β-SI |
| Akazawa et al. [30] | Japan | 26 (15) | - | Echo | No | Distensibility, β-SI |
| Singh et al. [31] | U.S | 15 (36.9) | 10 (42.9) | MRI | No | β-SI |
| Merlocco et al. [32] | U.S | 26 (25.11) | - | MRI | No | Distensibility, β-SI |
| Grillo et al. [33] | Italy | 51 (12) | 80 (11.9) | Echo | Yes | PWV |
| Salvi et al. [34] | Italy | 116 (33.7) | - | Echo | Yes | PWV |
| Selamet Tierney et al. [35] | U.S | 608 (11.2) | - | Echo | No | Distensibility, β-SI |
| Schäfer et al. [36] | U.S | 20 (18) | 22 (15) | MRI | Yes | PWV, Distensibility |
| Guala et al. [37] | Spain | 117 (25.3) | - | MRI | Yes | Distensibility |
| Yan et al. [38] | Germany | 69 (34.43) | 90 (67.27) | Echo | Yes | Distensibility, β-SI |
| Guala et al. [39] | Spain | 44 (36.95) | 36 (39.40) | MRI | Yes | PWV, Distensibility |
| Cui et al. [40] | Canada | 49 (17.9) | 87 (18.20) | Echo | Yes | PWV, β-SI |
| Andel et al. [41] | Netherlands | 35 (28) | - | MRI | Yes | Distensibility |
| Weismann et al. [42] | Sweden | 20 (22) | 67 (25) | Echo | Yes | PWV, Distensibility, β-SI |
| Cox et al. [43] | U.S | 32 (21.1) | - | Echo | Yes | Distensibility, β-SI |

In total, the review included data from 1925 patients with MFS and 836 patients without MFS, treated as controls, with mean age of participants ranging from 2 to 90 years old. A considerable fraction of the papers (10 out of 30) exclusively reported data for the Marfan cohort and did not include control data. As shown on Figure 2, diameter was primarily reported at the root and ascending aorta, with only about a third of the studies comparing Marfan and control groups. In contrast, PWV was predominantly reported in the ascending part of the aorta, with only one study that considered controls in the descending aorta (Dao) and another in the abdominal aorta (Abao). Distensibility was mostly reported in the ascending aorta, but only about a third of the papers included control data for comparison. The *β*-stiffness index was mainly reported in the root and ascending aorta.

**Figure 2:**
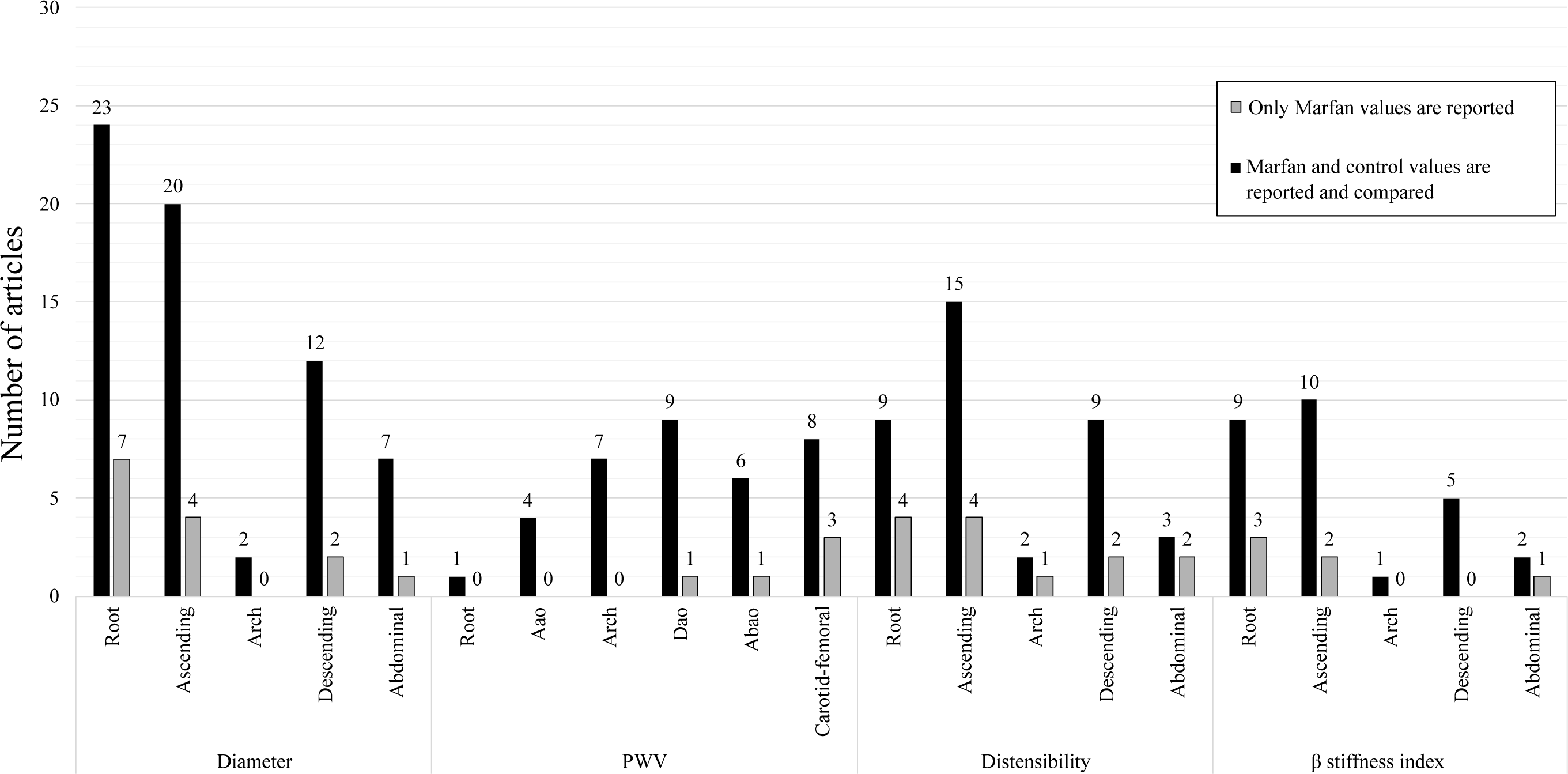
Number of articles reporting diameter, PWV, distensibility and *β* stiffness index at various locations. Black bars represent papers reporting only Marfan data, and gray bars represent papers including a control cohort and comparing results to Marfan.

In Figure 3, the size of the Marfan cohort is depicted in relation to their age. The horizontal bars displayed represent mean ± SD for each paper. Most of the articles encompassed cohorts with less than 60 patients, with age ranges spanning from pediatrics to 50 years old. Only three studies exceeded 100 participants and none of them included controls in their analysis.

**Figure 3:**
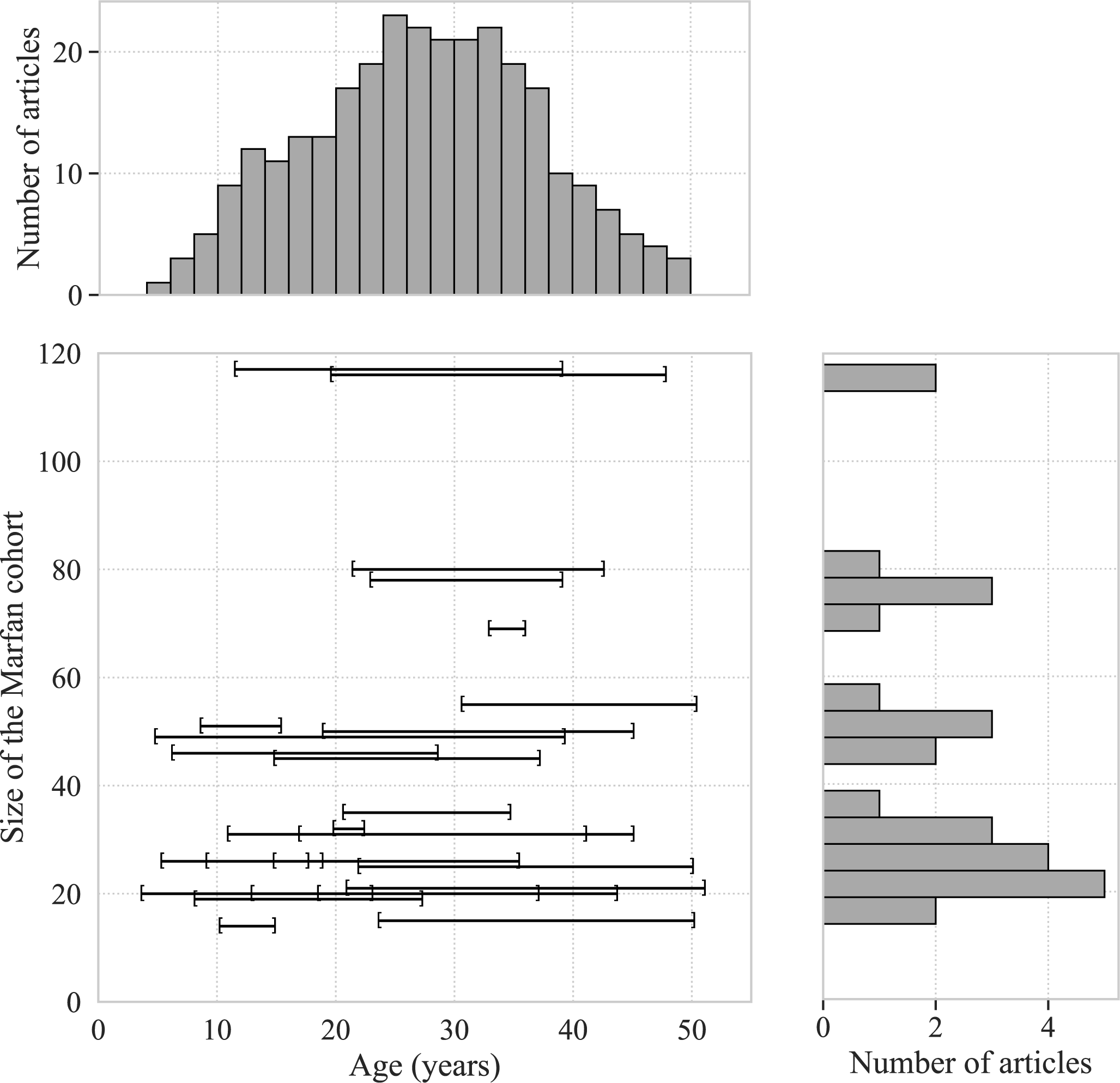
Mean age and standard deviation of the Marfan cohort in selected articles, organized by cohort size. The histogram on the top illustrates the number of articles containing patients within specific age ranges. The histogram on the right depicts the number of articles containing specific cohort sizes. One study (Selamet Tierney et al. [35]) is not included in this plot and consists of 608 MFS patients with a mean age of 11.2 and a standard deviation of 6.3.

### 3.3 Main findings from the selected papers

#### 3.3.1 Aortic diameter is the standard measure to compare Marfan and control

Among the 30 selected papers, 14 compared the diameter at the root between Marfan and control cohorts, 14 at the ascending aorta, and 3 at the descending aorta. Table 3 summarizes the results of these comparisons. The findings from these studies indicated significant differences in aortic size between the two groups. Notably, all papers reported a larger aortic root in Marfan patients, irrespective of the age of the cohort or correction for Body-Surface-Area (BSA). 10 studies found that the ascending aorta (Aao) was larger in Marfan patients, and 5 studies found no statistical difference in the diameters of the descending aorta (Dao).

**Table 3:**
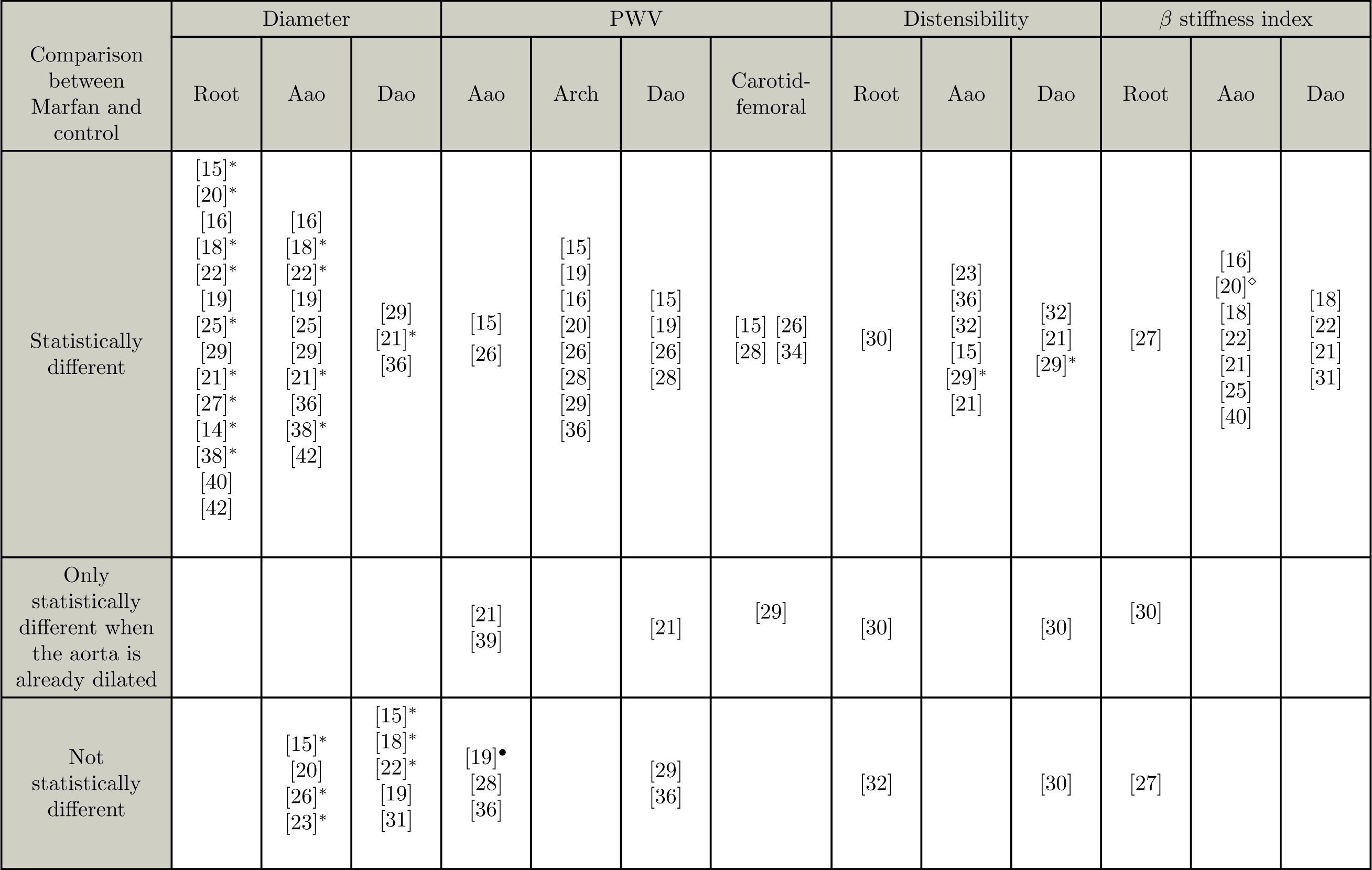
Studies comparing diameter, PWV, distensibility and *β*-SI between Marfan and control, and results of the statistical tests at different aortic locations. The symbol *∗* indicates quantities corrected for age, pulse pressure and diastolic area, ⋄ indicates that *β*-SI is corrected for sex, height and age, and • indicates that PWV is corrected for age and diameter.

#### 3.3.2 PWV is higher in Marfan patients, but only with dilated aortas

Among the selected studies, 7 compared PWV at the Aao between Marfan and control cohorts, 8 examined PWV at the arch, 7 in the descending aorta, and 5 from carotid to femoral (Table 3). Consistently, a significantly higher PWV was observed in all aortic regions from proximal to distal, as well as from carotid to femoral, in MFS patients. These differences remained valid even after adjusting for age and diameter. This finding highlights the presence of increased aortic stiffness in MFS patients across various regions of the aorta. However, in Oosterhof et al. [19] study, after correcting for age and diameter for the ascending aorta, no significant difference in PWV was observed between adult patients with and without Marfan Syndrome.

Additionally, in the studies conducted by Vitarelli et al. [21], Teixido-Tura et al. [29], and Guala et al. [39], it was observed that when separating the cohorts into dilated aortas and normal diameters, Marfan patients exhibited increased PWV compared to controls. However, this difference was significant only for already dilated aortas. According to Teixido-Tura et al. [29], compared with distensibility, PWV demonstrated a slower decrease at the early stage of aortic dilatation. This suggests that PWV might not be as sensitive to changes in aortic biomechanics during the initial stages of aortic dilatation compared to distensibility. However, as the aortic dilatation progresses, PWV gradually increases and eventually becomes significantly different between Marfan and control cohorts, but only when the aorta is already dilated.

#### 3.3.3 Aortic distensibility is lower in Marfan patients

Among the selected papers, 2 studies compared the distensibility at the root between Marfan and control cohorts, 7 studies examined the distensibility at the Aao, and 3 at the Dao (Table 3). Statistically lower distensibility was consistently reported in the aortic root, Aao, and Dao for Marfan patients. Furthermore, distensibility showed a significant decrease with age: Groenink et al. [15] highlighted that, compared to juvenile Marfan cohorts in the literature, the mean distensibility was nearly half in adult patients. Distensibility appears to be an early marker of biomechanical changes in Marfan Syndrome. In Akazawa et al. [30]’s study on children, a difference in distensibility of the root was observed between Marfan and control groups, even in non-dilated roots, at an early stage of life. In the Dao, distensibility did not differ whether the aorta was dilated or not.

Finally, studies conducted by Teixido-Tura et al. [29] and Vitarelli et al. [21] on older cohorts also reported lower distensibility in adult Marfan patients, whether their aortic root, ascending, or descending aortas were dilated or non-dilated. These results indicate a decrease in distensibility starting from the proximal aorta.

#### 3.3.4 ***β***-stiffness index is higher in Marfan patients

Among the selected studies, 2 compared the *β*-SI at the root between Marfan and control cohorts, 8 examined the *β*-SI at the Aao, and 4 at the Dao (Table 3). The findings consistently revealed that the *β*-SI was significantly higher in the root, Aao, and Dao in the MFS group when compared to control patients, even after adjusting for factors such as sex, age, and height. Notably, unlike PWV, MFS patients demonstrated higher *β*-SI values than controls in both cases of aortic dilatation and normal aortic diameters. However, it is essential to consider the observation by Wit et al. [27] that, after 40 years of age, the *β*-SI did not show significant differences between Marfan and control cohorts. This may have implications in understanding the progression of stiffness changes associated with age in Marfan Syndrome patients.

#### 3.3.5 Biomechanical stiffness measures can be early predictors of aortic dilatation

In the 17 longitudinal studies, Marfan patients were followed at various time points throughout their lives, and the evolution of stiffness parameters was measured. Statistical tests could identify a potential trend in the parameters’ evolution with age. The predictive power of each parameters was thus assessed with regards to aortic dilatation.

## PWV

In the study conducted by Groenink et al. [15], PWV was strongly correlated with age in control subjects at all levels of the aorta. However, in MFS patients, the increase in PWV with age was significantly higher in the proximal aorta compared to healthy subjects, supporting the hypothesis of media degradation starting at the root [34]. Despite its correlation with age, PWV was not found to be associated with progressive aortic dilatation at any level in the longitudinal study by Nollen et al. [17]. This suggests that PWV may not be a reliable candidate for predicting future aortic dilatation in MFS patients. Furthermore, in patients with aortic root replacement, even though the distensibility of the graft was significantly lower than the distensibility of the native aorta, the PWV showed no differences [17]. This indicates that PWV, as a regional measure, may not adequately differentiate diseased tissue locally and may be insensitive to differences in tissue composition.

A noteworthy exception is that PWV demonstrated high specificity and low sensitivity for predicting the absence of regional dilatation in MFS patients in the longitudinal study by Kröner et al. [28]. Specifically, at least 78% of MFS patients who showed no aortic growth at follow-up did not have increased regional PWV at baseline. Conversely, less than 33% of patients who presented with increased PWV at baseline had increased aortic growth at follow-up.

## Distensibility

Several studies, including Baumgartner et al. [18], Akazawa et al. [30], and Schäfer et al. [36], have demonstrated that distensibility can serve as a diagnostic parameter in addition to the current diameter measurements. The main reason is that distensibility was found to be lower even in patients with normal diameters at the root and ascending aorta. In a longitudinal study by Baumgartner et al. [22], the probability of developing an aneurysm was calculated based on ascending aortic distensibility. The findings revealed that higher distensibility measured at baseline was associated with a lower probability of developing aortic dilatation at follow-up. Similarly, Nollen et al. [17] demonstrated that distensibility was predictive of progressive descending thoracic aortic dilatation. A reduction of one unit in distensibility was associated with a 4-fold increase in the risk of dilatation, independent of aortic diameter. However, distensibility was not found to be a significant predictor of dilatation at other aortic locations, as noted by Teixido-Tura et al. [29]. This might be attributed to the relatively advanced stage of aortic disease in that particular study group. Additionally, Merlocco et al. [32] found a linear correlation between distensibility and age, with a slightly higher decline with age compared to normal subjects.

## ***β***-stiffness index

The study by Cox et al. [43] provided important insights into the relationship between the *β*-stiffness index and aortic dilatation in Marfan patients. Their findings revealed that the *β*-stiffness index in the aortic root was positively correlated with the dilatation rate, indicating that higher *β* stiffness values were associated with a faster rate of aortic dilatation. Interestingly, the baseline aortic root dimension alone did not show a significant correlation with the dilatation rate. This highlights the potential of the *β*-stiffness index as an independent and predictive measure for assessing aortic dilatation in Marfan patients.

Additionally, the *β*-stiffness index was the least dependant on blood pressure variation, making it a robust indicator of aortic stiffness, in comparison to distensibility [44]. Indeed, in Wada et al. [45] study on 7 subjects, no correlation was found between *β*-stiffness index and mean blood pressure. In Sugawara et al. [46] study, *β*-stiffness index did not change significantly after decreasing the blood pressure using *α*-adrenergic blockade.

### 3.4 Meta analysis

#### 3.4.1 Analysis of the augmented dataset

Through the data extraction and augmentation process, the dataset included 286 data points for diameter, 1063 for PWV, 1063 for distensibility, and 733 for the *β*-stiffness index. Out of these, 1278 data points were associated with a corresponding age value: 1027 age points had corresponding PWV, distensibility and *β*-SI, 194 only had corresponding PWV and distensibility, and 57 only had corresponding diameter. Original data points constituted 36% of the entire dataset, with the remaining being part of the augmentation process. Specifically for diameter, 62 data points were mean values with 11 converted using Eqs. 4 and 5, and 222 individual points were extracted from manuscripts using WebPlotDigitizer. For PWV, 464 points were calculated using conversion formulas, 39 points were mean values with 8 using Eqs. 4 and 5, and 544 individual data points were from the manuscripts. As for distensibility, 839 points were calculated using conversion formulas, 20 points were mean values with 9 using Eqs. 4 and 5, and 188 were individual data points. Lastly, for the *β*-stiffness index, 579 points were calculated using conversion formulas, 24 were mean values with 6 using Eqs. 4 and 5, and 238 were individual data points.

Out of the 1172 data points in the augmented dataset, the majority (1082) were associated with patients who did not undergo surgery, 90 data points were unknown (not specified in the article). Regarding medication, 139 data points were from patients under medication such as beta-blockers, 866 data points were from patients not under medication, and 167 data points were unknown (not specified in the article). Table 4 summarizes the number of data points in the augmented dataset obtained from each article.

**Table 4:** Papers included the meta-analysis and the number of datapoints in the augmented dataset.

| Papers | Number of datapoints |
| --- | --- |
| Wit et al. [27] | 244 |
| Westenberg et al. [26] | 149 |
| Cui et al. [40] | 134 |
| Teixido-Tura et al. [29] | 127 |
| Groenink et al. [15] | 109 |
| Oosterhof et al. [19] | 101 |
| Baumgartner et al. [18] | 95 |
| Weismann et al. [42] | 67 |
| Schäfer et al. [36] | 27 |
| Fattori et al. [23] | 19 |
| Baumgartner et al. [22] | 14 |
| Kiotsekoglou et al. [25] | 8 |
| Vitarelli et al. [21] | 8 |
| Bradley et al. [20] | 8 |
| Guala et al. [47] | 8 |
| Yan et al. [38] | 8 |
| Sandor et al. [16] | 6 |
| Kröner et al. [28] | 5 |
| Andel et al. [41] | 5 |
| Nollen et al. [17] | 5 |
| Salvi et al. [34] | 4 |
| Mortensen et al. [24] | 4 |
| Grillo et al. [33] | 4 |
| Guala et al. [37] | 2 |
| Cox et al. [43] | 1 |

#### 3.4.2 Comparison tests between Marfan and control on the augmented dataset

Consistent with previous literature findings, diameter was indeed statistically larger in Marfan patients at the Root (Figure 4), but no difference in the Aao and Dao was detected. Specifically, the mean diameter in the Marfan cohort was 3.91cm at the root, 2.98cm at the Aao, and 2.10cm in the Dao, compared to 3.05cm, 3.00cm, and 1.86cm, respectively, for the control cohort. Distensibility was significantly lower in Marfan patients except at the root after Bonferroni correction with mean values (in 10*^−^*^3^mmHg*^−^*^1^) of 2.46 in the Root, 3.57 in the Aao, 6.18 in the Arch, and 4.23 in the Dao, compared to 2.99 6.11, 8.17, and 6.56, respectively, for controls (Fig. 5). The *β*-SI was also higher in Marfan patients at all locations, with mean values of 13.93 in the root, 5.97 in the Aao, 4.42 in the Arch, and 6.53 in the Dao, compared to 8.25, 3.64, 3.36, and 4.90, respectively, for the control cohort (Fig. 5). Interestingly, no statistically different PWV was found in the Root and from carotid to femoral (Fig. 5). Table 5 summarizes the results of the statistical tests.

**Figure 4:**
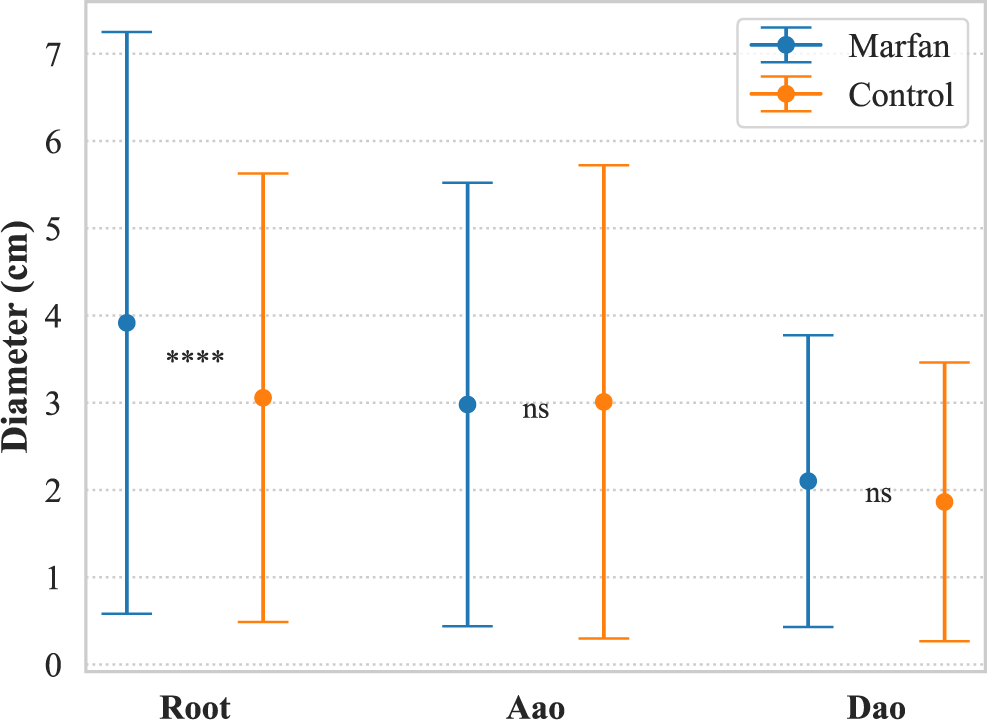
Mean and SD of aortic diameters in Marfan and control cohorts at three different locations show a signficant difference only at the root.

**Figure 5:**
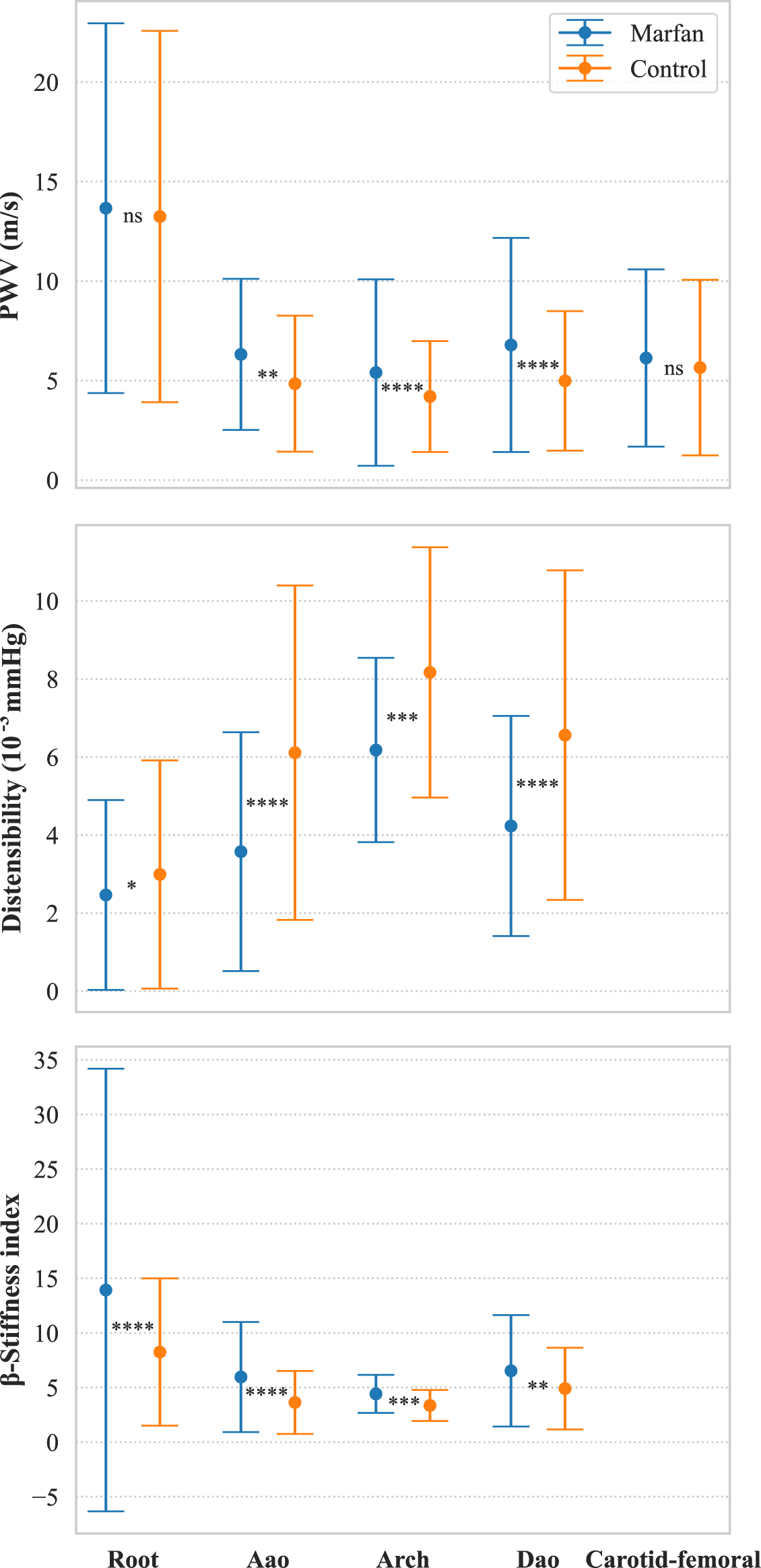
Mean and SD of Distensibility, PWV and *β*-stiffness index for the augmented dataset, without age consideration.

**Table 5:**
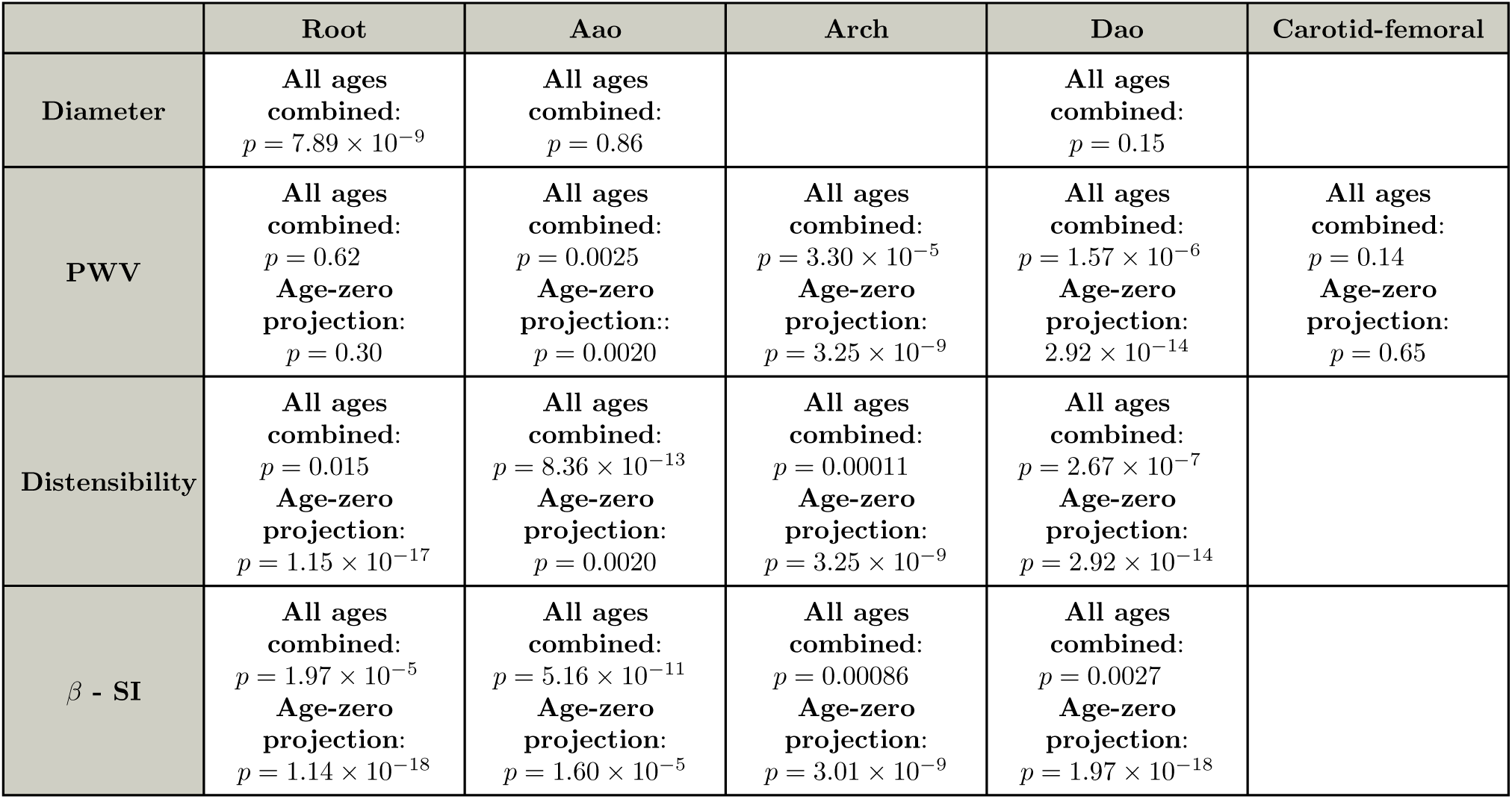
*p*-values for the Welch comparison tests between Marfan and control patients, for the three stiffness measures at different locations.

|  | Root | Aao | Arch | Dao | Carotid-femoral |
| --- | --- | --- | --- | --- | --- |
| Diameter | All ages combined:<br>$p = 7.89 \times 10^{-9}$ | All ages combined:<br>$p = 0.86$ | | All ages combined:<br>$p = 0.15$ | |
| PWV | All ages combined:<br>$p = 0.62$<br>Age-zero projection:<br>$p = 0.30$ | All ages combined:<br>$p = 0.0025$<br>Age-zero projection:<br>$p = 0.0020$ | All ages combined:<br>$p = 3.30 \times 10^{-5}$<br>Age-zero projection:<br>$p = 3.25 \times 10^{-9}$ | All ages combined:<br>$p = 1.57 \times 10^{-6}$<br>Age-zero projection:<br>$p = 2.92 \times 10^{-14}$ | All ages combined:<br>$p = 0.14$<br>Age-zero projection:<br>$p = 0.65$ |
| Distensibility | All ages combined:<br>$p = 0.015$<br>Age-zero projection:<br>$p = 1.15 \times 10^{-17}$ | All ages combined:<br>$p = 8.36 \times 10^{-13}$<br>Age-zero projection:<br>$p = 0.0020$ | All ages combined:<br>$p = 0.00011$<br>Age-zero projection:<br>$p = 3.25 \times 10^{-9}$ | All ages combined:<br>$p = 2.67 \times 10^{-7}$<br>Age-zero projection:<br>$p = 2.92 \times 10^{-14}$ | |
| $\beta$ - SI | All ages combined:<br>$p = 1.97 \times 10^{-5}$<br>Age-zero projection:<br>$p = 1.14 \times 10^{-18}$ | All ages combined:<br>$p = 5.16 \times 10^{-11}$<br>Age-zero projection:<br>$p = 1.60 \times 10^{-5}$ | All ages combined:<br>$p = 0.00086$<br>Age-zero projection:<br>$p = 3.01 \times 10^{-9}$ | All ages combined:<br>$p = 0.0027$<br>Age-zero projection:<br>$p = 1.97 \times 10^{-18}$ | |

#### 3.4.3 Linear regressions with age and projection at age-zero comparison

Figure 6 and Table 6 present the outcomes of the linear regressions at various aortic locations. Because the quantities were not normally distributed, the linear regressions were performed after log transformation. Figure 7 and Table 5 provides the results from the Welch comparison test after projecting the data to age-zero. The correlation analyses and age-zero projection tests revealed important findings regarding the relationship between PWV, distensibility, *β*-stiffness index, age, and Marfan syndrome.

**Figure 6:**
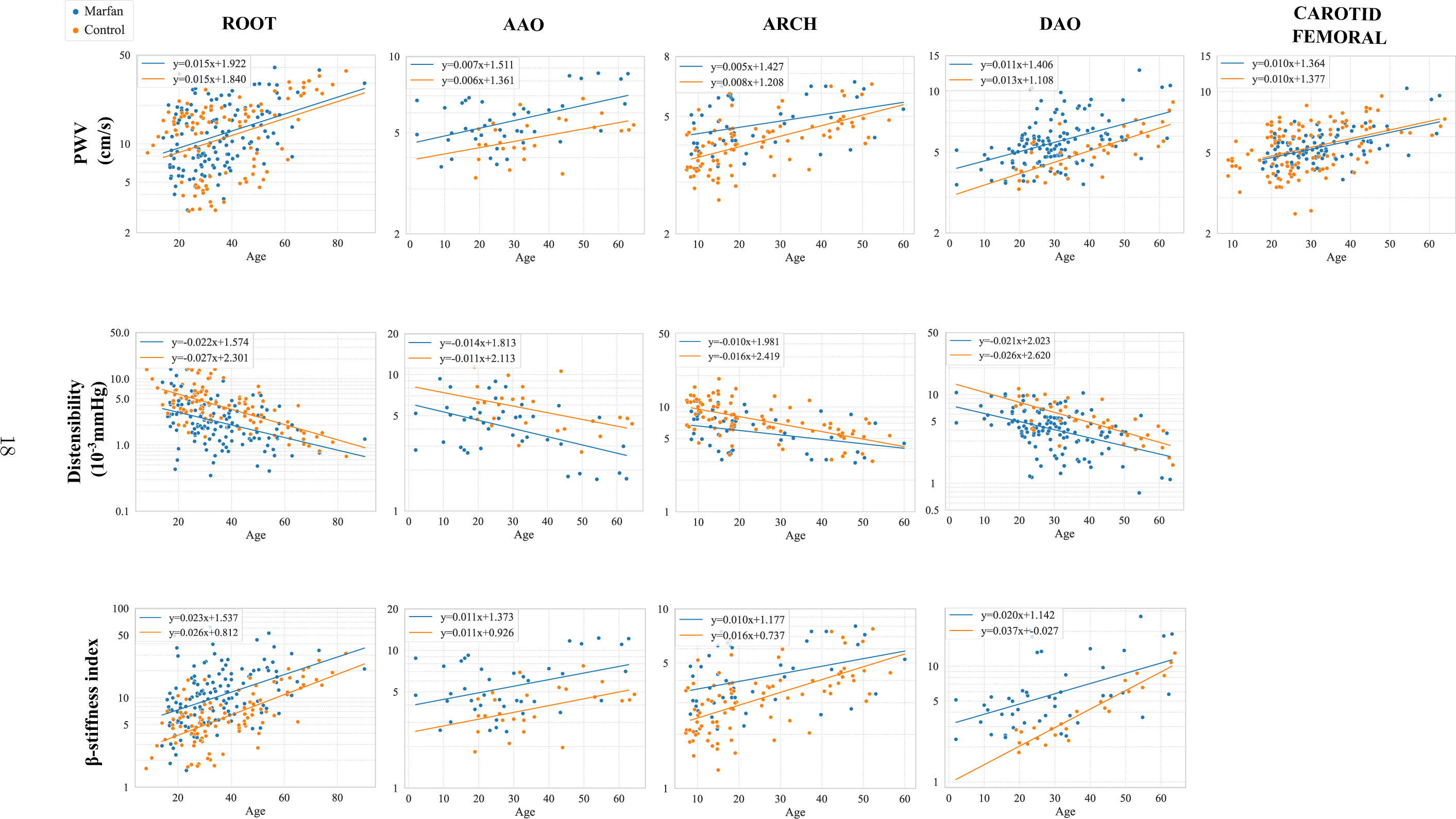
Linear regressions of PWV, distensibility and *β* stiffness index with respect to age at the different locations. Note the log scale on the vertical axis, since the regressions were performed on log-transformed values for each quantity to ensure normality.

**Figure 7:**
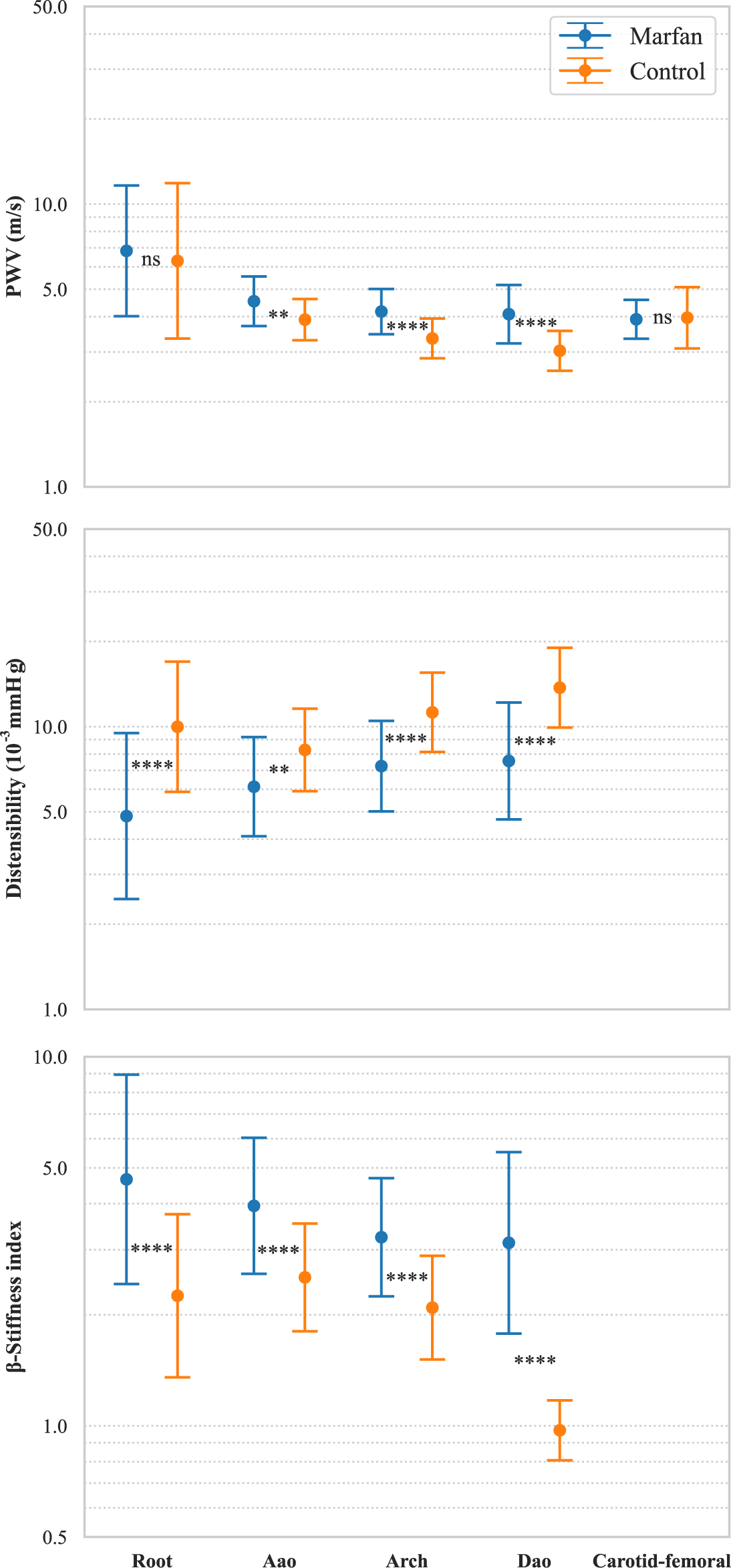
Mean and SD in log-scale of distensibility, PWV and *β*-stiffness index projected at zero age. The statistical tests were run on log-transformed values for each quantity.

**Table 6:** Results of the linear regression equations.

|  | Root | Aao | Arch | Dao | Carotid-femoral |
| --- | --- | --- | --- | --- | --- |
| PWV | **Marfan:**<br>$y = 0.015x + 1.92$<br>$R^2 = 0.14$ ;<br>$p = 1.11 \times 10^{-5}$<br>**Control:**<br>$y = 0.015x + 1.84$<br>$R^2 = 0.15$ ;<br>$p = 6.65 \times 10^{-5}$ | **Marfan:**<br>$y = 0.007x + 1.51$<br>$R^2 = 0.24$ ;<br>$p = 1.41 \times 10^{-3}$<br>**Control:**<br>$y = 0.0057x + 1.36$<br>$R^2 = 0.19$ ;<br>$p = 2.77 \times 10^{-2}$ | **Marfan:**<br>$y = 0.0049x + 1.43$<br>$R^2 = 0.13$ ;<br>$p = 1.70 \times 10^{-2}$<br>**Control:**<br>$y = 0.0082x + 1.21$<br>$R^2 = 0.35$ ;<br>$p = 2.81 \times 10^{-9}$ | **Marfan:**<br>$y = 0.011x + 1.41$<br>$R^2 = 0.20$ ;<br>$p = 2.04 \times 10^{-7}$<br>**Control:**<br>$y = 0.013x + 1.11$<br>$R^2 = 0.56$ ;<br>$p = 3.12 \times 10^{-8}$ | **Marfan:**<br>$y = 0.0095x + 1.36$<br>$R^2 = 0.26$ ;<br>$p = 5.17 \times 10^{-8}$<br>**Control:**<br>$y = 0.0098x + 1.38$<br>$R^2 = 0.18$ ;<br>$p = 4.78 \times 10^{-6}$ |
| Distensibility | **Marfan:**<br>$y = -0.022x + 1.57$<br>$R^2 = 0.17$ ;<br>$p = 9.25 \times 10^{-7}$<br>**Control:**<br>$y = -0.027x + 2.30$<br>$R^2 = 0.42$ ;<br>$p = 1.26e - 13$ | **Marfan:**<br>$y = -0.014x + 1.81$<br>$R^2 = 0.24$ ;<br>$p = 1.41 \times 10^{-3}$<br>**Control:**<br>$y = -0.011x + 2.11$<br>$R^2 = 0.19$ ;<br>$p = 2.77 \times 10^{-2}$ | **Marfan:** $y =$<br>$-0.0098x + 1.98$<br>$R^2 = 0.13$ ;<br>$p = 1.70 \times 10^{-2}$<br>**Control:**<br>$y = -0.016x + 2.42$<br>$R^2 = 0.34$ ;<br>$p = 2.81 \times 10^{-9}$ | **Marfan:** $y =$<br>$-0.021x + 2.023$<br>$R^2 = 0.20$ ;<br>$p = 2.04 \times 10^{-7}$<br>**Control:**<br>$y = -0.026x + 2.62$<br>$R^2 = 0.56$ ;<br>$p = 3.12 \times 10^{-8}$ | |
| $\beta$ - SI | **Marfan:**<br>$y = 0.022x + 1.54$<br>$R^2 = 0.19$ ;<br>$p = 1.81 \times 10^{-7}$<br>**Control:**<br>$y = 0.026x + 0.81$<br>$R^2 = 0.44$ ;<br>$p = 3.58 \times 10^{-14}$ | **Marfan:**<br>$y = 0.011x + 1.37$<br>$R^2 = 0.15$ ;<br>$p = 1.35 \times 10^{-2}$<br>**Control:**<br>$y = 0.011x + 0.93$<br>$R^2 = 0.19$ ;<br>$p = 2.77 \times 10^{-2}$ | **Marfan:**<br>$y = 0.0098x + 1.18$<br>$R^2 = 0.13$ ;<br>$p = 1.70 \times 10^{-2}$<br>**Control:**<br>$y = 0.016x + 0.74$<br>$R^2 = 0.34$ ;<br>$p = 2.81 \times 10^{-9}$ | **Marfan:**<br>$y = 0.020x + 1.14$<br>$R^2 = 0.24$ ;<br>$p = 5.64 \times 10^{-4}$<br>**Control:**<br>$y = 0.037x - 0.027$<br>$R^2 = 0.89$ ;<br>$p = 3.10 \times 10^{-10}$ | |

Notably, a positive correlation was observed between PWV and age in both Marfan and control patients at all aortic locations (*p <* 0.05). However, the age projection suggested that there was no statistically significant difference in PWV at the root (*p* = 0.30) and from carotid to femoral (*p* = 0.65) between Marfan and control patients at age-zero. Additionally, both MFS and control patients exhibited a negative correlation between distensibility and age at all aortic locations. The age-zero projection demonstrated that distensibility was lower, indicating higher aortic stiffness in MFS patients already at birth. However, the slopes of the linear regressions revealed that distensibility decreased more rapidly for the control cohort compared to Marfan patients (–0.027 vs. –0.022 for MFS in the root, –0.016 vs. –0.010 in the Arch, and –0.026 vs. –0.021 in the Dao, all values in 10*^−^*^3^mmHg*^−^*^1^ per year). This result could also be influenced by the fact that the meta-analysis does not include patients who have had surgery. Since patients who have had surgery tend to be older, this may introduce a bias in the dataset, potentially affecting the observed rate of distensibility decline in the Marfan cohort. The correlation analyses showed that *β*-SI was positively correlated with age in both Marfan and control patients at all aortic locations (*p <* 0.05). After conducting age-zero projection, the results confirmed that there is a statistically significant difference in *β*-stiffness index between Marfan and control patients at all locations. Marfan patients had higher values after age-zero projection, indicating stiffer tissues at an early stage of life compared to controls.

## 4 Discussion

### 4.1 Key findings in the literature

The diameter of blood vessels is a commonly used indicator for detecting biomechanical changes, and abnormal diameters at specific locations in the aorta are considered signs of disease. Diameter was reported to be larger in Marfan patients at the root and Aao. These results align with previously observed patterns in pediatric Marfan patients, where dilatation typically begins at the sinus of Valsalva, followed by the sino-tubular junction (STJ), and is less frequent in the descending aorta [48]. The reported higher diameters at the root in MFS patients also provide support for the current guideline of measuring the aortic root to plan prophylactic surgery.

Across most aortic regions, studies reported a significantly higher PWV in MFS when compared to control cohorts, but only in the already dilated aortas. Indeed, PWV was not as sensitive to changes in aortic biomechanics in the initial phases of aortic dilatation, e.g., compared to distensibility. This observation underscores the importance of considering the stage of aortic dilatation when interpreting PWV values and highlights the complexity of using PWV as a sole predictor for aortic dilatation in MFS patients. While it may provide valuable information about aortic stiffness, its ability to precisely predict aortic growth is limited. Combining PWV with other relevant parameters, such as distensibility, may yield more comprehensive insights into the dynamics of aortic changes in MFS and aid in making accurate clinical assessments.

The results derived from the studies included in this review provided evidence that change in distensibility values can serve as an effective mean to detect alterations in the biomechanical properties of the aorta. Specifically, a decrease in distensibility was indicative of increased tissue stiffness within the aortic wall. Distensibility was lower at all aortic locations even in juvenile non-dilated aortas, with values half of those in the healthy patients. A significant correlation between distensibility and age was also put forward. Finally, longitudinal studies showed that a higher distensibility is associated with higher chances of developing an aneurysm in the Aao and Dao. Although sensitive to location, these findings underscore the potential utility of distensibility as an early indicator of aortic biomechanical changes in Marfan patients.

The results across the studies indicated that *β*-SI values were notably higher in the root, Aao, and Dao among patients with Marfan Syndrome in comparison to control subjects, even in non-dilated aortas. It was positively correlated with the rate of aortic dilatation, emphasizing its role as a valuable indicator. Unlike baseline aortic dimensions, *β*-SI appeared to be a better predictor of the progression of aortic dilatation. However, it is worth noting that, after 40 years of age, the control and MFS cohorts could not be distinguished using the *β*-SI. Interestingly, younger MFS patients had higher *β*-SI values compared to their age-matched controls, but this distinction diminished in older populations. This particular observation offers insights on progression of aortic stiffness in MFS patients as they age. It may suggest that altered aortic stiffness eventually leads to convergence between the Marfan and control cohorts after a certain age threshold.

### 4.2 Meta-analysis results

In the meta-analysis, in consensus with the literature, diameter was found to be significantly higher in MFS at the root, which align with the current guideline of measuring the aortic root to plan prophylactic surgery. Interestingly, no difference was found in the Aao and Dao. In this case, however, no age consideration was possible to due lack of data reporting diameter with respect to age. Combining all patients regardless of their age includes bias when comparing Marfan and control since diameter depends highly on age, weight and height of the patient. Diameter assessment adjusted for body surface area, which has been found to be more useful than age, height, or weight alone for the measuring the size of the aorta [49], would have possibly led to different statistical results.

The meta-analysis results also provide valuable insights into the relationship between aortic stiffness measures, age, and Marfan syndrome. Firstly, it was observed that without considering age, Marfan patients exhibited higher PWV values at all locations, except for the root and the carotid-femoral region. It is particularly interesting to note that distinctions between cohorts were challenging when assessing PWV at the carotid-femoral region, which is the most commonly used PWV measurement in clinical practice. This observation can be explained by the fact that carotid-femoral PWV covers a substantial portion of the aortic tract and may not be sensitive to local variations in tissue stiffness. On the other hand, PWV was found to be approximately two times higher at the root. However, the root’s relatively small length may lead to difficulties in tracking the foot of the pulse wave, resulting in the considerably large standard deviation observed in patients. PWV at the root is rarely utilized in clinical practice and failed to distinguish between the two cohorts effectively. Furthermore, a positive correlation was identified between PWV and age in both Marfan and control patients across all aortic locations. However, the age-zero projection suggested that a statistically significant difference in PWV exists at all locations, except at the root and the carotid-femoral region. This notable result implies that age plays a significant role in the evolution of PWV. More importantly, it indicates that the difference between Marfan and control patients does not develop with age. Instead, Marfan patients are born with higher PWV, indicating stiffer aortic tissues. It is worth highlighting that PWV measurements taken at the carotid-femoral region and the root fail to capture these inherent differences.

Both Marfan and control patients were found to exhibit a negative correlation between distensibility and age at all aortic locations. The age-zero projection further emphasized that distensibility is lower, signifying increased aortic stiffness in MFS patients, even at a young age. Notably, the slopes of the linear regressions indicated that distensibility decreases at a faster rate for the control cohort compared to Marfan patients. It is essential to note that this observation might be influenced by the absence of patients past 40 years old in the dataset, introducing a potential bias.

Additionally, our analysis demonstrated that *β*-stiffness index was positively correlated with age in both Marfan and control patients at all aortic locations. Before and after conducting age-zero projection, the results affirmed a statistically significant difference in *β*-stiffness index between Marfan and control patients at all locations. These findings suggest that Marfan patients exhibit higher *β*-SI values even at an early stage of life, indicating stiffer aortic tissues compared to controls.

The results at age-zero projection provide a more reliable assessment of aortic stiffness in Marfan patients and underscore the importance of considering age as a confounding factor in such studies. Overall, the data indicates that distensibility and *β*-index are consistently altered in Marfan patients compared to controls, while PWV still shows location-specific differences between the two groups before and after age-zero projection.

### 4.3 Recommendations for future studies

Handling missing data poses a significant challenge in systematic quantitative reviews. This review underscores the importance of reporting data to facilitate statistically robust and comprehensive meta-analyses. Rather than reporting meand and SD, we recommend that studies report individual data points. This could be achieved through supplementary dataset if needed. The incorporation of a substantial number of participants and the diverse range of study designs greatly contributes to the robustness and depth of the findings presented in this review. However, if each patient had reported diameter, age and stiffness measure, it would have allowed for a more thorough statistical analysis (such as multivariate regression).

It is noticeable that there is a lack of studies focusing on specific age ranges and cohorts with more than 40 patients. For pediatrics, only one multicenter study comprises 608 patients [35]. A plausible explantation could be that MFS is rarely diagnosed in paediatric populations since patients do not appear phenotypically different. Not all paediatric patients have family history eihter, which makes the diagnosis at an early stage of life challenging. Similarly, no studies focused on patients past 40 years old, which represents a turning point in stiffness increase according to Wit et al. [27]. This absence could potentially be attributed to the fact that aortic surgeries are typically performed before patients reach this age, resulting in limited accessible data for older patients.

### 4.4 Limitations

Regarding limitations, we acknowledge that all pertinent studies may not have been captured in our search. Our search criteria might have missed studies that were not explicitly categorized under, or did not explicitly reference a Marfan syndrome diagnosis. Since Marfan syndrome can be misdiagnosed for other connective tissue disorders caused by pathogenic variants in genes other than FBN1, studies published before the revised Ghent criteria were excluded. However, they likely include true Marfan patients and provide valuable insights. Similarly, mild Marfan syndrome cases and cases of suspected but not yet verified Marfan syndrome, were possibly overlooked in this present work. Despite the revised Ghent criteria, Marfan syndrome clinically overlaps with other connective tissue disorders, such as Loeys-Dietz syndrome, and distinguishing them is challenging in the absence of a molecular diagnosis. Consequently, individuals with mutations in proteins related to the TGF-*β* pathway might receive a Marfan syndrome diagnosis against the Ghent nosology and be included in this study.

While this work does touch upon aortic diameter, it is important to note that a significant portion of the existing literature primarily focuses on aortic diameter in Marfan patients, but these studies were not encompassed in this review. Instead, discussions concerning diameter in this review are derived exclusively from data within the selected papers that primarily address aortic stiffness measures.

The aortic sites and regions were not defined identically between the selected papers. To facilitate the reporting of results and minimize potential inconsistencies, we made efforts to categorize them into five main regions: Root, Aao, Arch, Dao and Carotid-femoral, although minor variations may remain. The data gap in specific age brackets is an essential consideration when interpreting the findings of this review. The review’s strength lies in its ability to compile and analyze aortic stiffness in different populations. However, the absence of age-specific studies, especially in the pediatric and older age groups, highlights a potential area for future research.

Concerning the meta-analysis, despite our efforts to address missing data, several factors introduced bias into the statistical tests. This bias stems from theoretical conversion equations, which cannot precisely mimic physiological behaviour, as well as the absence of individual data points and the lack of age-associated values for each data point. Similarly, conducting an age projection involves making inferences beyond the data range, which may not align with in-vivo behaviour. The regression analysis with age was conducted solely on data that included individual data points with corresponding age. Additionally, the predictive power of aortic stiffness measures could not be thoroughly investigated in the meta-analysis due to the absence of articles reporting individual values for patients followed at various ages. Lastly, examining diameter as a potential confounding variable was unfeasible because of the scarcity of studies reporting individual data points for stiffness measures, alongside corresponding age and diameter values.

### 4.5 Conclusion

To the best of the authors’ knowledge, this is the first systematic review and meta analysis investigating how in-vivo aortic stiffness measures can be early markers of aortic disease in Marfan syndrome, and their prediction of aortic dilatation. Our study emphasizes the importance of using a combination of parameters, including diameter measurements and stiffness indices, to obtain a more comprehensive evaluation of aortic disease in MFS patients. This approach can provide a deeper understanding of disease progression and assist clinical decision making.

## Funding

This study was partially funded by the Engineering and Physical Sciences Research Council (EP/N02124X/1) and University of Glasgow’s College of Science and Engineering via PhD studentship.

## Data availability statement

All data produced in the present study are available upon reasonable request to the authors

## Notes

### Competing Interest Statement

The authors have declared no competing interest.

